# Unlocking the potential of wearable device wear time to enhance postpartum depression screening and detection

**DOI:** 10.1101/2024.10.07.24315026

**Authors:** Eric Hurwitz, Samantha Meltzer-Brody, Zachary Butzin-Dozier, Rena C. Patel, Noémie Elhadad, Melissa A. Haendel

## Abstract

Postpartum depression (PPD) is a mood disorder affecting one in seven women after childbirth that is often under-screened and under-detected. If not diagnosed and treated, PPD is associated with long-term developmental challenges in the child and maternal morbidity. Wearable technologies, such as smartwatches and fitness trackers (e.g., Fitbit), offer continuous and longitudinal digital phenotyping for mood disorder diagnosis and monitoring, with device wear time being an important yet understudied aspect. Using the *All of Us* Research Program (AoURP) dataset, we assessed the percentage of days women with PPD wore Fitbit devices across pre-pregnancy, pregnancy, postpartum, and PPD periods, as determined by electronic health records. Wear time was compared in women with and without PPD using linear regression models. Results showed a strong trend that women in the PPD cohort wore their Fitbits more those without PPD during the postpartum (PPD: mean=72.9%, SE=13.8%; non-PPD: mean=58.9%, SE=12.2%, *P*-value=0.09) and PPD time periods (PPD: mean=70.7%, SE=14.5%; non-PPD: mean=55.6%, SE=12.9%, *P*-value=0.08). We hypothesize this may be attributed to hypervigilance, given the common co-occurrence of anxiety symptoms among women with PPD. Future studies should assess the link between PPD, hypervigilance, and wear time patterns. We envision that device wear patterns with digital biomarkers like sleep and physical activity could enhance early PPD detection using machine learning by alerting clinicians to potential concerns facilitating timely screenings, which may have implications for other mental health disorders.

## Introduction

The rise of wearable device ownership, such as Fitbits®, have led to significant advancements in the realm of digital phenotyping^1^. Because wearables can be used to monitor the same individual in a continuous and longitudinal manner, their use for personalized medicine is exciting, especially for mental health where individualized tools for diagnosis and treatment monitoring are lacking. Digital biomarkers from wearables are collected in a passive manner in non-clinical settings, thus enabling these devices to offer potential enhancements to several clinical aspects of the mental health care continuum^2^.

Postpartum depression (PPD) is a mood disorder that is one of the most common complications of childbirth^3^. PPD has significant implications for maternal morbidity, associations with developmental delays for the child, and incurs significant costs to society^4–7^. Because PPD is a highly heterogeneous condition and often stigmatized, many patients go undiagnosed^8^. One significant issue with PPD is that most women do not receive sufficient screening, as only about 31% of women with PPD receive a diagnosis^4^. As noted by Cox et al., there are reliable screening instruments for PPD (e.g., the Edinburgh-Postnatal Depression Scale [EPDS])^4,9^ and specific treatments for PPD (e.g., brexanalone and zuranolone)^10,11^; yet screening and diagnosis of PPD lag behind and novel approaches for PPD detection are direly needed.

Wearables have provided an opportunistic route for enhanced behavioral phenotyping during pregnancy and in the postpartum period, including for PPD^2,12,13^. Given the under-diagnostic rate of PPD, readily available consumer wearables may aid in its early detection due to their rise in ownership and passive data collection, thereby improving patient outcomes. For example, our recent work demonstrated that individualized machine learning (ML) models using digital biomarkers (heart rate, physical activity, and energy expenditure) from a Fitbit were able to distinguish between four phases of pregnancy, including pre-pregnancy, pregnancy, postpartum, and during PPD^12^. Wearable devices have also been shown to predict whether a woman will experience preterm birth using only one week of activity and sleep data^13^. Additionally, studies have demonstrated that activity intensity distribution during the day, resting heart rate, and heart rate variability captured from a wearable device were predictive of maternal loneliness, which is associated with PPD^14^. Collectively, these studies highlight a relationship between digital biomarkers and perinatal mental health, suggesting that wearables may enhance longitudinal monitoring.

While it has been shown that digital biomarkers from wearables, such as the Fitbit, combined with ML can provide insight into mental health conditions, patterns of wear time remain relatively unexplored. Prior studies exploring wearable device wear time have mainly taken place in the human-computer interaction field in a general population and disease-agnostic setting^15–19^. A few studies have looked at wear time behavior in the context of biomedical research, but only in a limited capacity. For instance, analyses from the Framingham Heart Study suggest that higher depressive symptoms are associated with lower smartwatch use, defined as wearing the device for more than five hours at least one day of the week. The authors suggest this observation is due to the link between motivation and depressive symptoms, where individuals are less likely to engage with a smartwatch for health-related activities like tracking daily steps or promoting healthy lifestyle behaviors^20^. While this result posits a relationship between device wear time and mental health, there is a need to explore wearable device wear time in pregnancy cohorts, which are heterogeneous and constantly changing, making it difficult to identify potential screening tools and biomarkers.

In this study, we sought to demonstrate the value of wearable device wear time as an insightful digital biomarker in digital mental health. We leveraged the *All of Us* Research Program dataset (AoURP), a longitudinal dataset with several health-related data types, including as electronic health records (EHRs), surveys, physical measurements, and Fitbit data^21^. To highlight the potential value of Fitbit wear time in facilitating early detection of PPD, we characterized differences in wear time between women with and without PPD. We propose that wearable device wear time may serve as a clinically informative biomarker to help facilitate early detection of mental health disorders in a continuous, passive, and non-clinical setting. For PPD specifically, gaining insight into wearable behavior patterns could offer valuable understanding of perinatal mental health, potentially enhancing screening and diagnosis in real-world settings.

## Methods

### Data sources and platforms

Data in this study leveraged the AoURP Controlled Tier v7 data set. Analysis was conducted using the AoURP Researcher Workbench cloud platform. All phenotyping and data analysis were conducted using R. Fitbit data in the AoURP operates under a bring-your-own-device model, where participants who consent to participate in the study share data from their device that they already own^22^.

### Ethical considerations

The protocol for the AoURP study was reviewed by the institutional review board of the AoURP (protocol 2021-02-TN-001). The institutional review board follows the regulations and guidance of the National Institutes of Health Office for Human Research Protections for all studies, ensuring that the rights and welfare of research participants are overseen and protected uniformly. The informed consent process states that participants have the option to withdraw at any time. Privacy of participant data is maintained in the following three ways: 1) storing data on protected computers, 2) preventing researchers from seeing identifiable patient information, such as name or social security number, 3) having researchers sign a contract they won’t try to identify participants. Furthermore, access to the AoURP dataset is only available through the Researcher Workbench, which is only to researchers who have completed the requisite training at institutions with a signed Data Use Agreement. For compensation, participants are offered $25 one-time in the form of cash, gift card, or an electronic voucher if they are asked and decide to go to an *All of Us* partner center for physical measurements to give blood, saliva, or urine samples. Of note, other racial/ethnic groups were not reported since the sample sizes for several of them were less than 20 and could risk patient reidentification, which violates the AoURP Dissemination policy^23^.

### Computational phenotyping of PPD and non-PPD cohorts

Women were assigned to the PPD cohort using the same approach that we described previously^12,24^. Briefly, identifying women with PPD consists of a three-fold approach: 1) a PPD diagnosis, 2) a diagnosis of depression during the postpartum period, or 3) antidepressant drug exposure during the postpartum period. Women were assigned to the non-PPD cohort by identifying women with available pregnancy or delivery EHR data in a similar manner to the PPD cohort and then excluding those who were in the PPD cohort.

To assess wear time behavior in a longitudinal manner, Fitbit wear time data for each woman in the PPD cohort was assigned to one of four time periods: 1) pre-pregnancy (starting from two years prior to the PPD index date), 2) pregnancy, 3) postpartum without depression (after the delivery date and before the PPD diagnosis date), or 4) PPD (a diagnosis up to 24 months from the date of delivery, which has been done in prior work and in this study also represents a time period)^25,26^. The PPD time period ranged from 14 days prior to the index date through 30 days after the index date, which was selected because 1) the diagnostic criteria for PPD requires that women display 5 depressive symptoms lasting 2 weeks and 2) some individuals received antidepressant medication on the same date as their index date, which can take effect after 4 weeks of use^27,28^.

Because women in the non-PPD group didn’t undergo a “fourth” phase of PPD as seen in the PPD group, we introduced a pseudo-time period called the *PPD-equivalent* phase as a time frame for the non-PPD group to align with the PPD phase. The index date for the PPD-equivalent period was set at 58 days following delivery, corresponding to the median number of days after delivery of PPD diagnosis among women in the PPD group, following the same strategy we employed in our prior work. Similarly, 14 days prior to the index date was not used since these women did not actually experience PPD^12^. Women were only included in the PPD or non-PPD cohorts if they had any Fitbit data during any of the four time periods.

### Measuring and comparing Fitbit wear time between PPD and non-PPD cohorts

Fitbit wear time was measured by first determining the number of hours a smartwatch was worn in a day using methods described previously^29^. Prior studies have indicated that a “valid” day of smartwatch data requires 10 hours of wear time and between 100 and 45000 steps. In this study, rather than analyze days of “valid” data, we wanted to understand patterns of Fitbit wear time behavior amongst women with PPD. Hence, we established a binary variable for each day to indicate whether the device was worn or not based on the presence of at least 1 hour of wear time, where hours of wear time were measured based on the presence of step data, similar to prior studies^29^. We then determined the percentage of days the Fitbit was worn during each of the four time periods (i.e., pre-pregnancy, pregnancy, postpartum, and PPD [or PPD-equivalent for the non-PPD cohort]) by counting the number of days the device was worn divided by the total number of days during that time period for each woman. The total number of days was determined for each person by filtering data after the first recorded date of any Fitbit data to ensure that we were not labeling someone as not wearing their smartwatch if they didn’t even own one. Fitbit wear time was compared between PPD and non-PPD cohorts using linear regression, where four total models were run (one for each time period). Each model filtered on data during one time period and the means were calculated using the emmeans() function^30^. Models were run with covariates of age at PPD diagnosis (or age at the index date for the non-PPD cohort) and race/ethnicity at a significance level of 0.05.

### Measuring and comparing the number of hours Fitbit devices were worn per day between PPD and non-PPD cohorts

We determined the number of hours per day the Fitbit was worn using the same logic as described above. To assess whether there was a difference in the number of daily hours the Fitbit was worn between PPD and non-PPD cohorts during each time period, we ran a linear mixed effects model using the lme4 package in R since there were multiple days of data per person (i.e., person ID was included as the random effect) at a significance level of 0.05^31,32^. The dataset was filtered on individuals who had at least 1 hour of wear time, since we wanted to ensure that we were assessing whether there was a difference in the number of hours the device was worn per day among days that the device was actually worn. Models were also run adjusting for age at PPD diagnosis (or age at the index date for the non-PPD cohort) and race/ethnicity at a significance level of 0.05.

### Measuring and comparing the percentage of days Fitbits were worn to sleep between PPD and non-PPD cohorts

In order to assess how often women with PPD wore their device to sleep, we focused on whether women had any record of “main sleep” for each date as a binary variable for yes or no. We then determined the percentage of days the Fitbit was worn to sleep during each of the four time periods (i.e., pre-pregnancy, pregnancy, postpartum, and PPD [or PPD-equivalent for the non-PPD cohort]) by counting the number of days the device was worn to sleep divided by the total number of days during that time period for each woman. The analysis was run in a similar fashion as described for comparing the percentage of days the device was worn, where a linear regression model was used to assess if there was a difference in the percentage of days the device was worn to sleep between women in the PPD and non-PPD cohorts during each of the time periods at a significance level of 0.05. Models were rerun with covariates of age at PPD diagnosis (or age at the index date for the non-PPD cohort) and race/ethnicity at a significance level of 0.05.

### Assessing the correlation between device wear time prior to and during PPD

The correlation between device wear time prior to PPD with device wear time during PPD was performed by filtering on two time periods of interest (e.g., pre-pregnancy and PPD) and assessing the correlation amongst all women in the PPD cohort. The ggpubr package was used to determine the correlation and was evaluated at a significance level of 0.05^33^. The same analysis was conducted comparing device usage during pregnancy and PPD time periods as well. Both analyses were repeated in women without PPD for comparison.

### Assessing the correlation between device consistency prior to and during PPD

Device wear time consistency was measured by determining the maximum number of consecutive days the Fitbit was worn for each time period for each unique person. Device wear time for one day was defined using the same definition as before, where we considered an individual wore the device if they had at least one hour of wear time for each date based on the presence of step data^29^. We then determined the relationship between device wear time consistency during pre-pregnancy and PPD by calculating the Pearson correlation coefficient between the max number of days the device was worn during the PPD time period versus the pre-pregnancy time period at a significance level of 0.05 using the ggpubr package in R^33,34^. We also performed the same analysis replacing the pre-pregnancy time period with the pregnancy time period to assess the correlation between device consistency during pregnancy and PPD. This process was repeated in the non-PPD cohort for comparison.

### Large language models

ChatGPT (GPT-3.5), developed by OpenAI (https://openai.com/) was used to edit some portions of the manuscript, including grammar, language, and synonyms. All recommendations from ChatGPT were reviewed by the author and were not used for the purpose of generating ideas or content.

## Results

### Descriptive statistics

Women were determined to be in the PPD cohort using methods as previously described (see Methods section; Figure 1)^12,24^. Our AoURP cohort consisted of 142 women who experienced pregnancy and had available Fitbit data with a total of 108062 days of data, where 41 women experienced PPD (31201 days of data) and 101 women (76861 days of data) did not. To achieve an accurate comparison in women without PPD, we created a pseudo-time period labeled *PPD-equivalent* starting 58 days following delivery, which was the median number of days after delivery for PPD diagnosis (see more details about pregnancy time periods in the Methods section). In the PPD and non-PPD cohorts, there were 13225 and 40212 days of data during pre-pregnancy, 11055 and 27559 days during pregnancy, 5089 and 6060 days during postpartum, and 1832 and 3030 days of data during the PPD/PPD-equivalent time periods, respectively. The median age of the PPD cohort was 33.1 years old (interquartile range [IQR]=29.1-35.7) compared to 33.9 years old (IQR=30.9-37.1) for women in the non-PPD cohort. Both the PPD (87.8%) and non-PPD cohorts (75.2%) were predominantly white non-Hispanic (Table 1).

**Figure 1:**
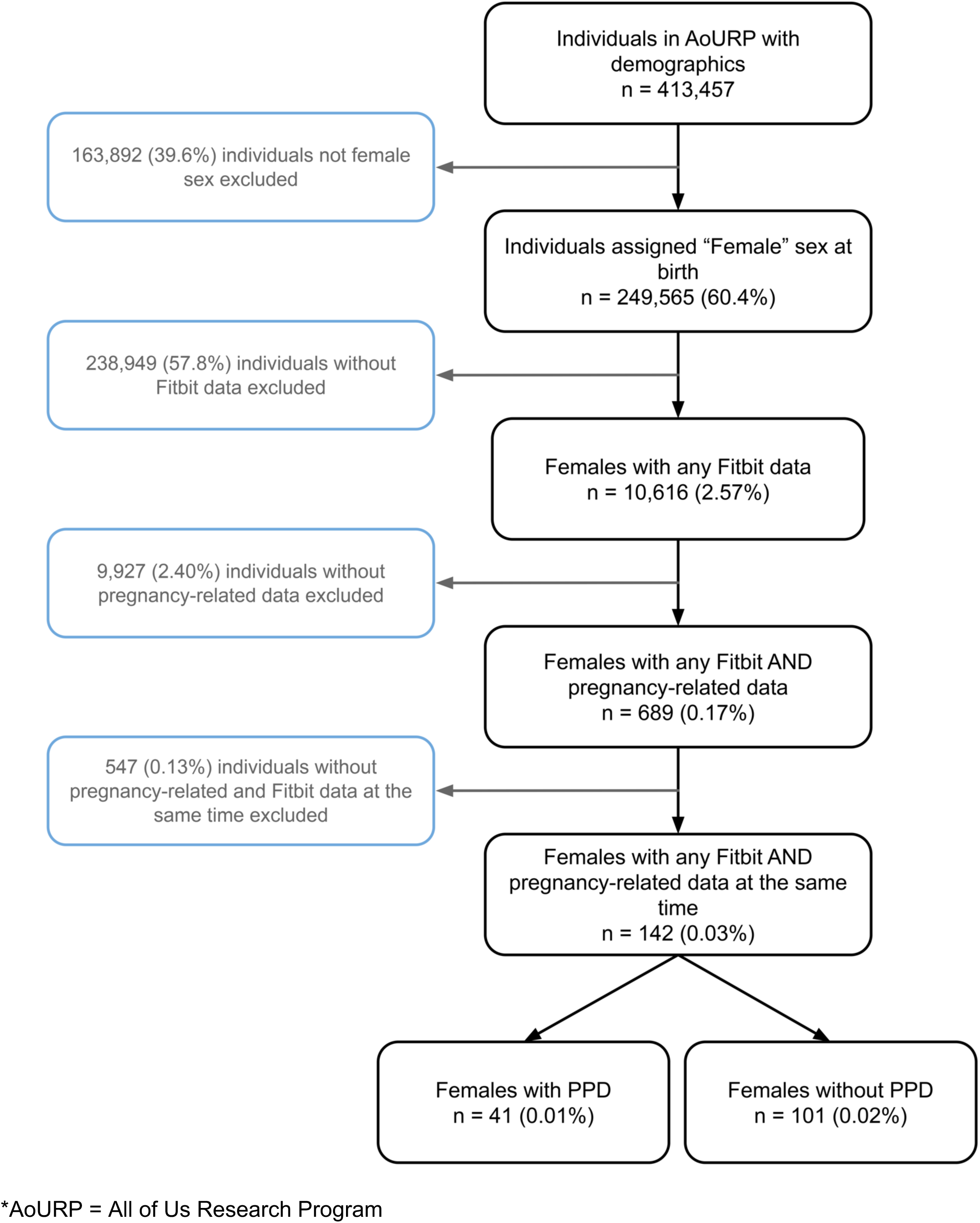
Flow diagram describing the inclusion and exclusion criteria of postpartum depression (PPD) and non PPD cohorts.

**Table 1:**
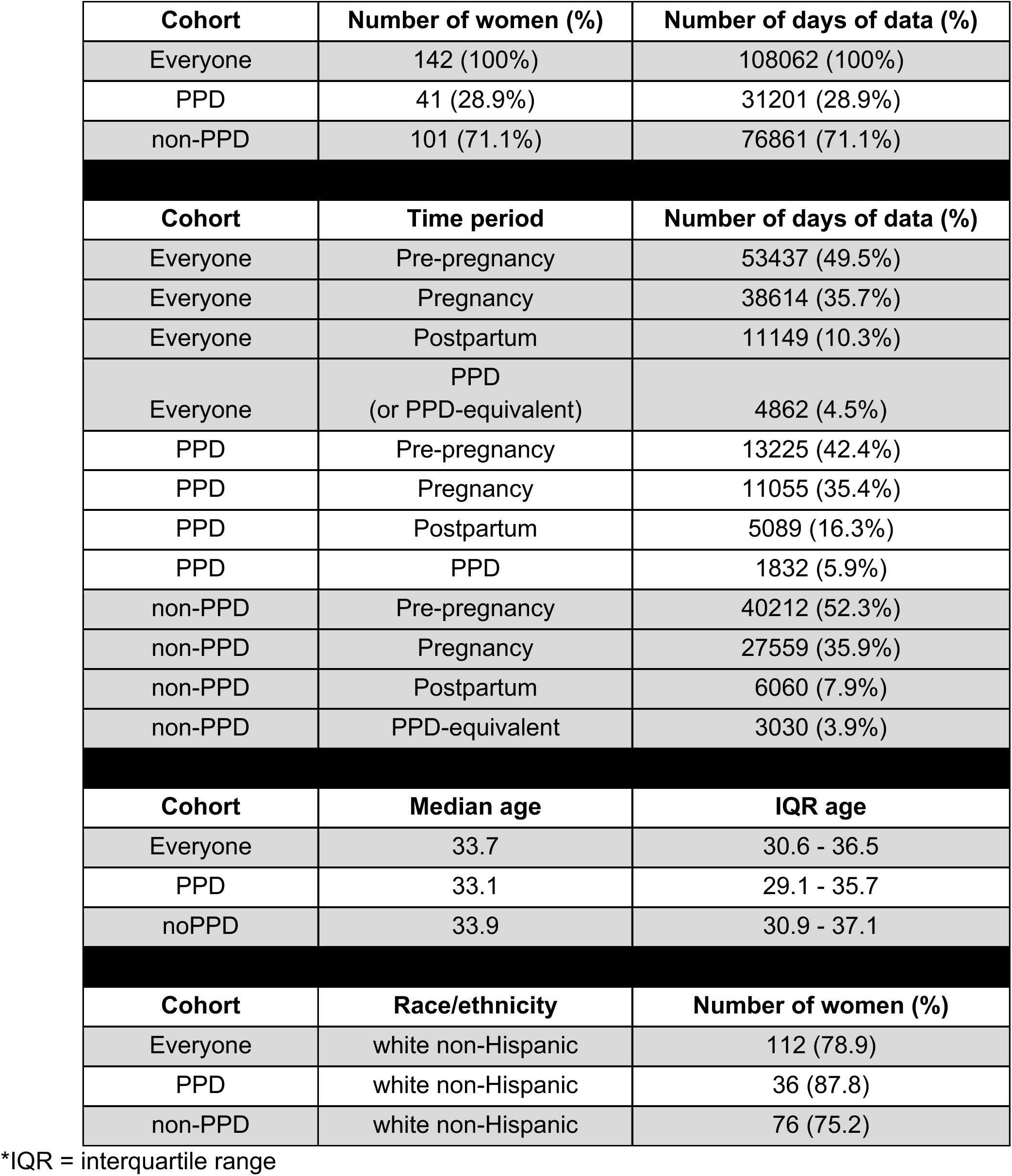
Descriptive statistics of women in the PPD and non-PPD cohorts.

| **Cohort** | **Number of women (%)** | **Number of days of data (%)** |
| --- | --- | --- |
| Everyone | 142 (100%) | 108062 (100%) |
| PPD | 41 (28.9%) | 31201 (28.9%) |
| non-PPD | 101 (71.1%) | 76861 (71.1%) |
| **Cohort** | **Time period** | **Number of days of data (%)** |
| Everyone | Pre-pregnancy | 53437 (49.5%) |
| Everyone | Pregnancy | 38614 (35.7%) |
| Everyone | Postpartum | 11149 (10.3%) |
| Everyone | PPD<br>(or PPD-equivalent) | 4862 (4.5%) |
| PPD | Pre-pregnancy | 13225 (42.4%) |
| PPD | Pregnancy | 11055 (35.4%) |
| PPD | Postpartum | 5089 (16.3%) |
| PPD | PPD | 1832 (5.9%) |
| non-PPD | Pre-pregnancy | 40212 (52.3%) |
| non-PPD | Pregnancy | 27559 (35.9%) |
| non-PPD | Postpartum | 6060 (7.9%) |
| non-PPD | PPD-equivalent | 3030 (3.9%) |
| **Cohort** | **Median age** | **IQR age** |
| Everyone | 33.7 | 30.6 - 36.5 |
| PPD | 33.1 | 29.1 - 35.7 |
| noPPD | 33.9 | 30.9 - 37.1 |
| **Cohort** | **Race/ethnicity** | **Number of women (%)** |
| Everyone | white non-Hispanic | 112 (78.9) |
| PPD | white non-Hispanic | 36 (87.8) |
| non-PPD | white non-Hispanic | 76 (75.2) |

\*IQR = interquartile range

### Women in the PPD cohort wore their devices more often during the postpartum and PPD periods compared to those in the non-PPD cohort

We first sought to evaluate whether women with PPD displayed Fitbit wear time behavior that differed to those without PPD. We calculated the percentage of days that each woman wore their device during the PPD and PPD-equivalent time periods and built a linear regression model adjusted for age at PPD diagnosis and race/ethnicity. The results revealed a strong trend that the percentage of days the device was worn in the PPD cohort (mean=70.7%, standard error [SE]=14.5%) was greater than the non-PPD cohort with a mean of 55.6% (SE=12.9%, *P*-value = 0.08; Figure 2).

**Figure 2:**
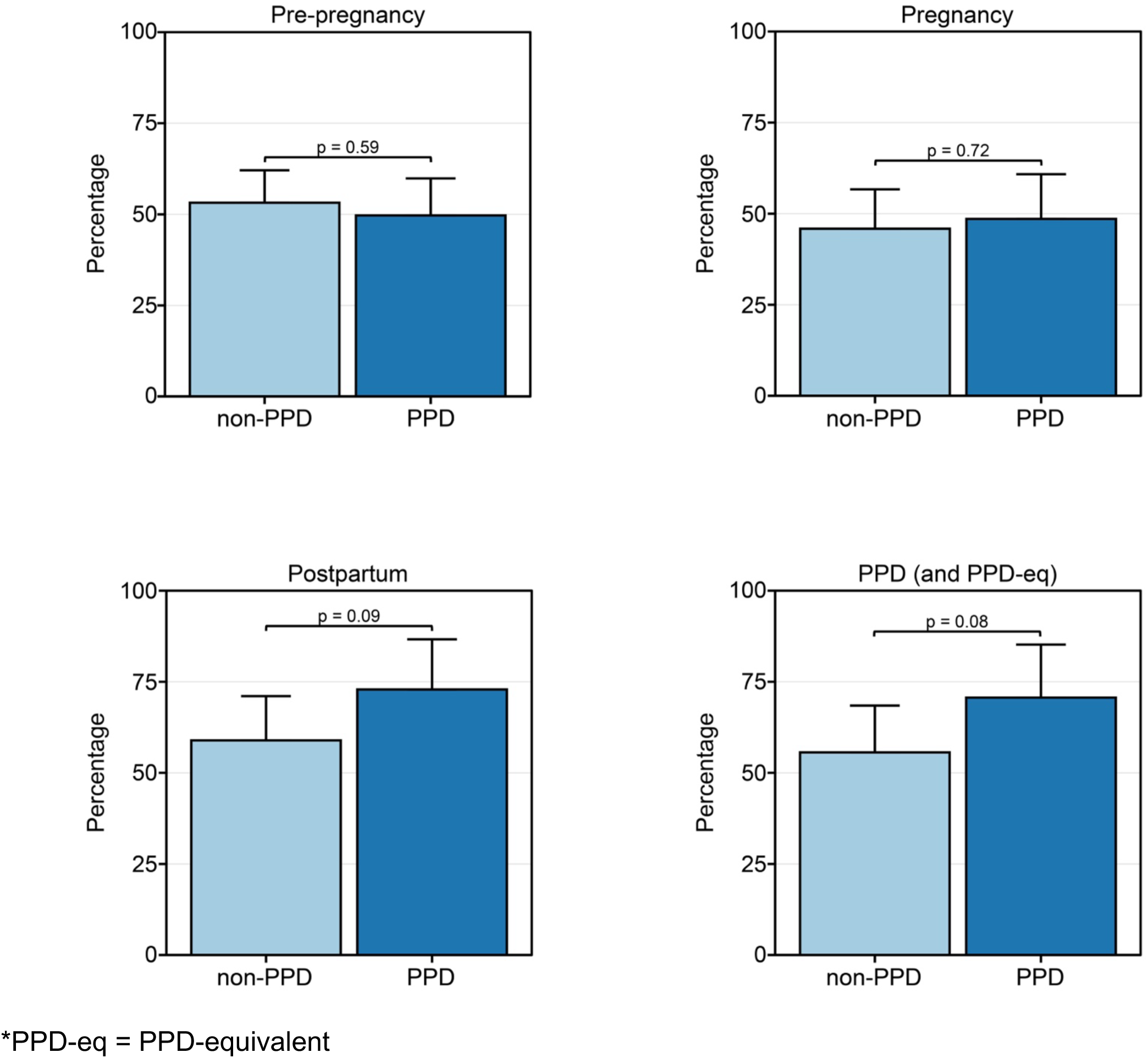
Women in the PPD cohort tended to wear their wearable device more than those in the non-PPD cohort during the postpartum and PPD time periods. The percentage of days women in the PPD and non-PPD cohorts wore their wearable device during the pre-pregnancy (top left), pregnancy (top right), postpartum (bottom left), and PPD (or PPD-equivalent; bottom right) time periods. Data in the PPD and non-PPD cohorts were compared using linear regression adjusted for age at PPD diagnosis and race/ethnicity and are expressed as mean ± standard error.

Observing this pattern during the PPD/PPD-equivalent time frames, we proceeded to explore potential disparities in wear time between PPD and non-PPD cohorts across other pregnancy stages, including pre-pregnancy, pregnancy, and postpartum periods. Such analysis aims to discern potential associations between Fitbit wear time behavior and future PPD onset. Models were run in a similar fashion for the pre-pregnancy, pregnancy, and postpartum time periods, where we also detected a strong trend of increased wear time during the postpartum time period, with a mean of 72.9% (SE=12.8%) in the PPD cohort compared to 58.9% (SE=12.2%) in the non-PPD cohort (*P*-value=0.09; Figure 2). These results suggest that women who go on to develop PPD may wear their device more than those who don’t in the postpartum period. Alternatively, there was no significant difference in the percentage of days the device was worn during pre-pregnancy or pregnancy time periods between the two cohorts (Figure 2).

### We did not detect a significant difference in the number of hours per day wearables were worn between PPD and non-PPD cohorts

Observing variation in the percentage of days wearable devices were worn between PPD and non-PPD cohorts, we subsequently evaluated if there were any differences in the daily duration of device wear time adjusted for age at PPD diagnosis and race/ethnicity. Surprisingly, our findings revealed no trends or significant differences between PPD and non-PPD cohorts during pre-pregnancy (PPD: mean=14.4, SE=0.98; non-PPD: mean=14.8, SE=0.87, *P*-value=0.50), pregnancy (PPD: mean=15.8, SE=0.98; non-PPD: mean=16.4, SE=0.85, *P*-value=0.36), postpartum (PPD: mean=16.5, SE=1.00; non-PPD: mean=17.2, SE=0.84, *P*-value=0.29), or the PPD/PPD-equivalent periods (PPD: mean=17.4, SE=0.97; non-PPD: mean=17.8, SE=0.84, *P*-value=0.53; Figure 3).

**Figure 3:**
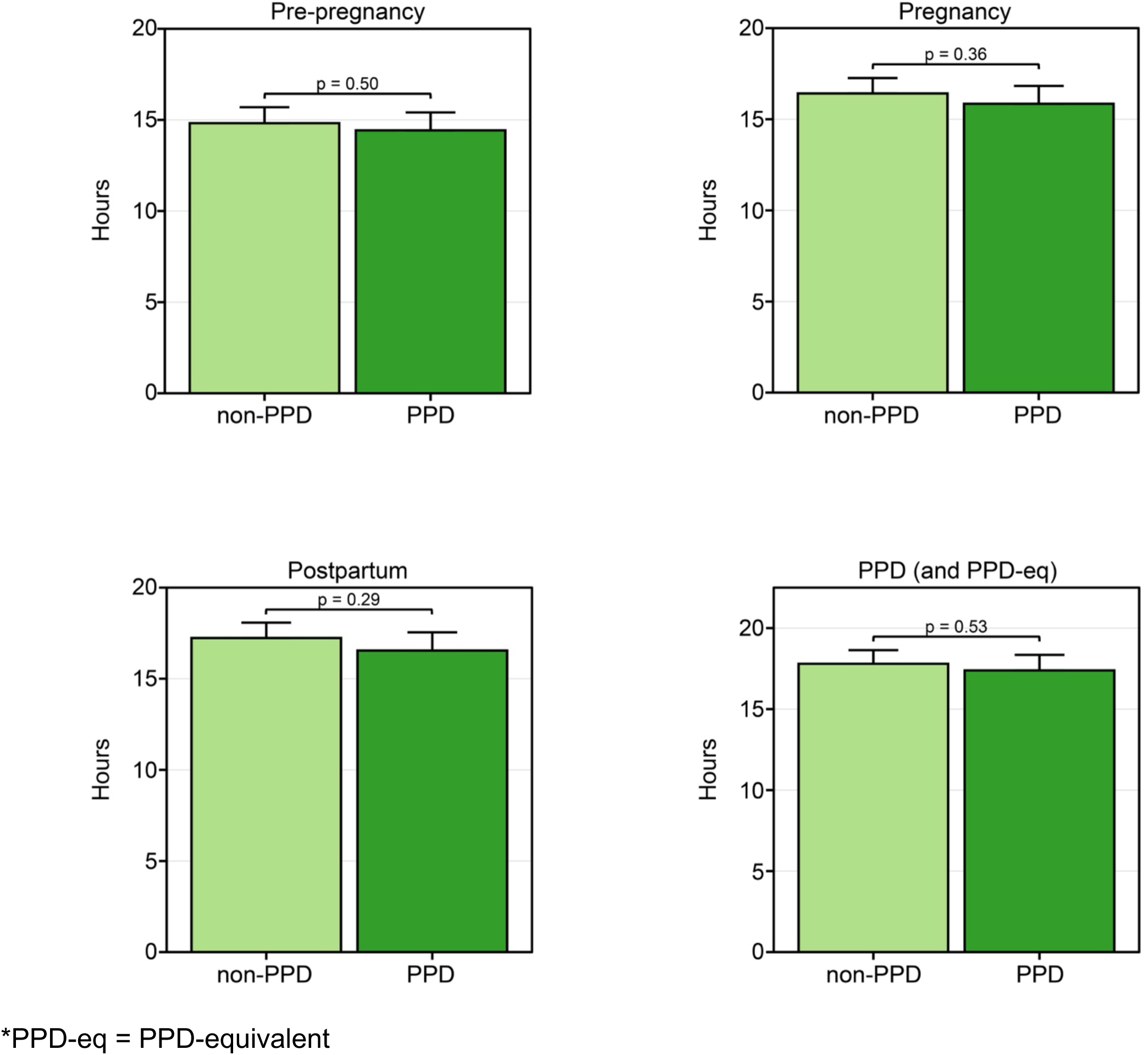
Women in the PPD and non-PPD cohorts wore their Fitbit for the same number of hours per day across time periods of pregnancy. The number of hours per day women in the PPD and non-PPD cohorts wore their wearable device during the pre-pregnancy (top left), pregnancy (top right), postpartum (bottom left), and PPD (or PPD-equivalent; bottom right) time periods. Data in the PPD and non-PPD cohorts were compared using linear mixed-effects models with person ID as the random effect adjusted for age at PPD diagnosis and race/ethnicity and are expressed as mean ± standard error.

### Women in the PPD cohort wore their Fitbit to sleep more during the postpartum and PPD periods compared to those without PPD

Given the extensive connection between sleep and PPD, we sought to describe and compare the percentage of days women in the PPD cohort wore their Fitbit to sleep compared to those without PPD during each phase of pregnancy^35–41^. When comparing the percentage of days women in each cohort wore their Fitbit to sleep, the results displayed a similar observation as the percentage of wear time, where we noticed a trend of women wearing the device to sleep more during the postpartum period in the PPD cohort (mean=58.9%, SE=12.2%) compared to the non-PPD cohort (mean=45.6%, SE=10.8%, *P*-value=0.07) adjusted for age at PPD diagnosis and race/ethnicity (Figure 4). There was a similar trend during the time period women experienced PPD (mean=64.4%, SE=13.4%) compared to those who did not (mean=49.8%, SE=11.8%, *P*-value=0.07; Figure 4). No differences were detected in the percentage of days Fitbits were worn to sleep between PPD and non-PPD cohorts during the pre-pregnancy or pregnancy time periods (Figure 4).

**Figure 4:**
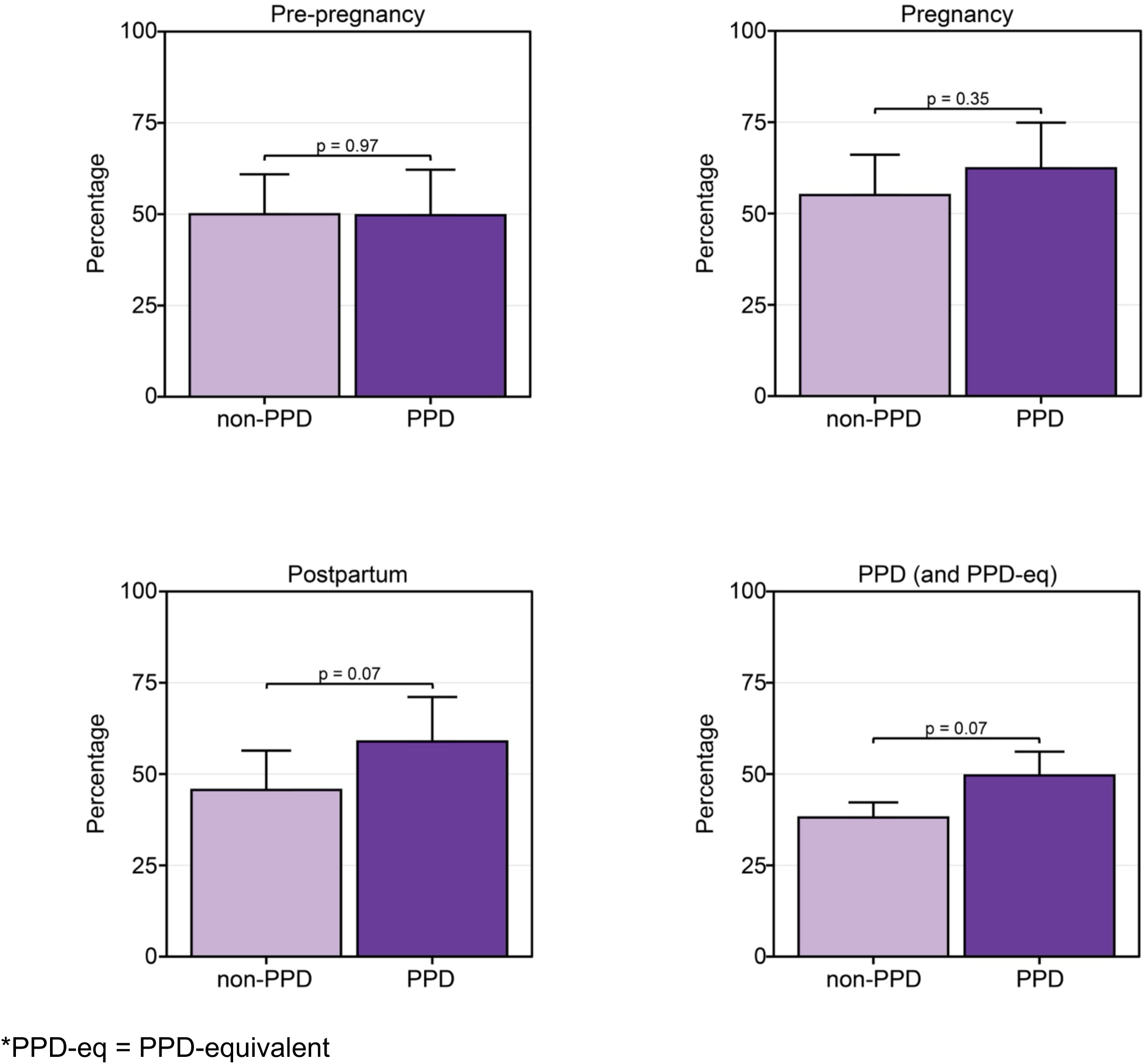
Women in the PPD cohort tended to wear their devices more during sleep than those in the non-PPD cohort during the postpartum and PPD time periods. The percentage of days women in the PPD and non-PPD cohorts wore their wearable device to sleep during the pre-pregnancy (top left), pregnancy (top right), postpartum (bottom left), and PPD (or PPD-equivalent; bottom right) time periods. Data in the PPD and non-PPD cohorts were compared using linear regression adjusted for age at PPD diagnosis and race/ethnicity and are expressed as mean ± standard error.

### Fitbit wear time consistency during pre-pregnancy was not correlated with consistency during PPD, but did correlate among women without PPD

Lastly, we wanted to explore individual-level device wear time patterns before and during PPD. Therefore, we examined the correlation between the percentage of days within women who wore their smartwatch during time periods prior to PPD (i.e., pre-pregnancy and pregnancy) with the PPD time period. For instance, a positive correlation would suggest that those who wore their smartwatch more frequently during the pre-pregnancy period tended to do so during the PPD time period. We conducted this analysis in parallel with the non-PPD cohort for comparison. In women with PPD, the results displayed a significant positive correlation between the percentage of days the Fitbit was worn during pre-pregnancy and PPD time periods (*r*=0.48, *P*-value=0.005; Figure 5). A positive correlation was also detected during pre-pregnancy and PPD-equivalent time periods among women without PPD, although it was not as strong (*r*=0.24, *P*-value=0.018; Figure 5). There also existed a strong positive correlation between the percentage of wear time during the pregnancy and PPD time periods among women in the PPD (*r*=0.77, *P*-value<0.001) and non-PPD cohorts (*r*=0.66, *P*-value<0.001; Figure 5).

**Figure 5:**
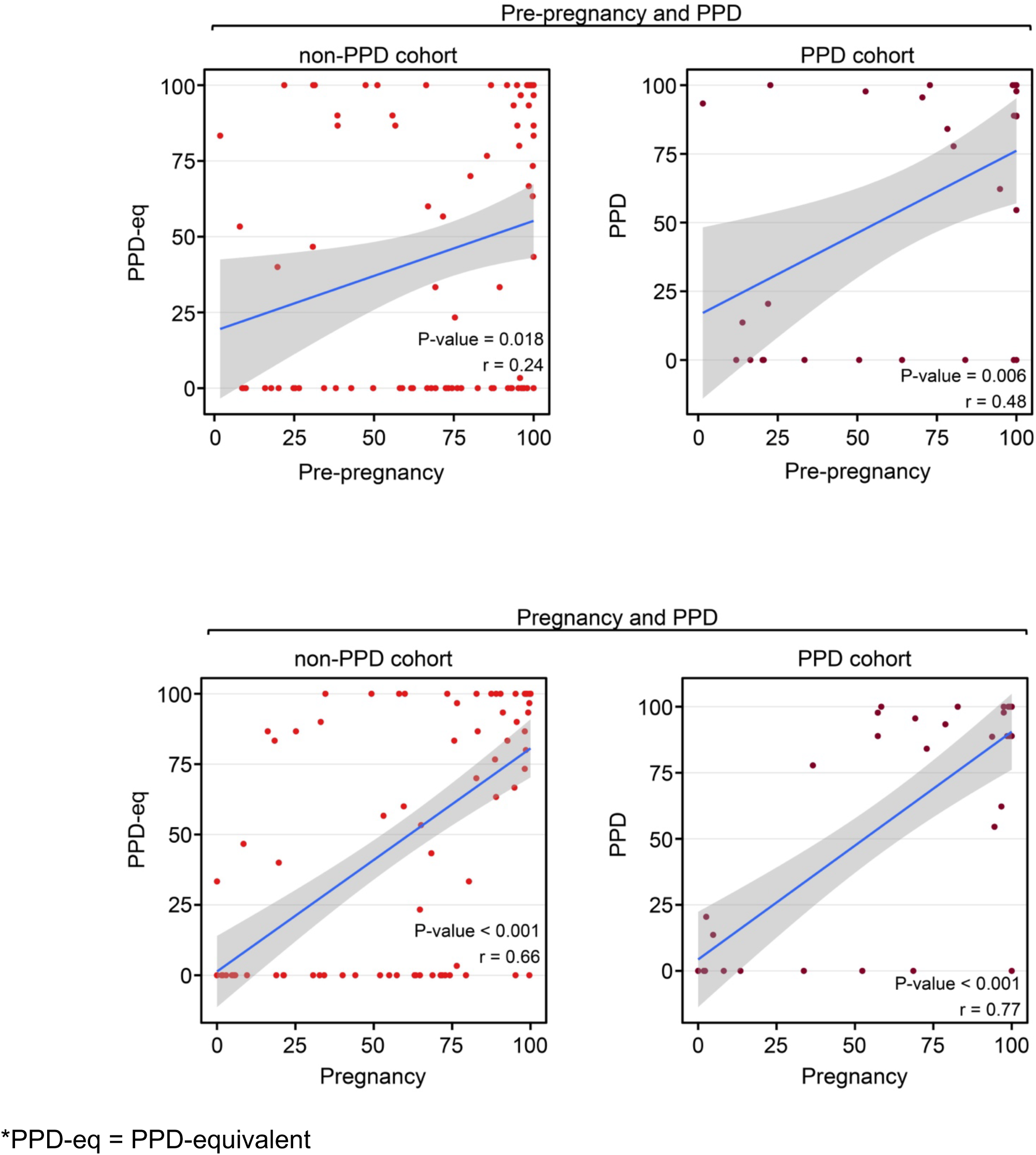
Women in the PPD and non-PPD cohorts displayed the same pattern of correlation between wear time during earlier and later pregnancy time periods. The Pearson correlation coefficient between the percentage of days women wore their Fitbit during pre-pregnancy and PPD (top) in addition to pregnancy and PPD (bottom). The blue line represents the line of best fit and gray shading shows the 95% confidence interval.

To further understand the wear time behavior of women and their Fitbit during each time period, we also sought to analyze the consistency with which the device was worn. Our earlier analyses focused on comparing the percentage of days the device was worn across different time periods; however, we acknowledged that wear patterns could vary. For example, if there were 50 days in total to potentially wear the device during one of the time periods, wearing it consistently for 25 consecutive days followed by 25 days of non-use is different from alternating between wearing and not wearing the device every other day, even though both scenarios indicate 50% wear time. Therefore, to assess individual-level consistency during each time period, we determined the maximum consecutive number of days the device was worn during each time period and examined the correlation across time periods (i.e., during pre-pregnancy and PPD [or PPD-equivalent]; during pregnancy and PPD [or PPD-equivalent]). The results displayed a strong trend in wear time consistency between pre-pregnancy and PPD-equivalent time periods among women without PPD (*r*=0.25, *P*-value=0.07), while those with PPD did not exhibit any correlation (*r*=-0.05, *P*-value=0.84; Figure 6). Alternatively, a significant correlation was present in both the PPD (*r*=0.48, *P*-value=0.02) and non-PPD (*r*=0.54, *P*-value<0.001) cohorts between the pregnancy and PPD (or PPD-equivalent) time periods (Figure 6). These data suggest a relationship between the consistency of Fitbit usage during pregnancy and the PPD (or PPD-equivalent) periods in both cohorts. Notably, when analyzing the consistency of smartwatch wear time during pre-pregnancy, the relationship only was present among women in the non-PPD cohort (Figure 6).

**Figure 6:**
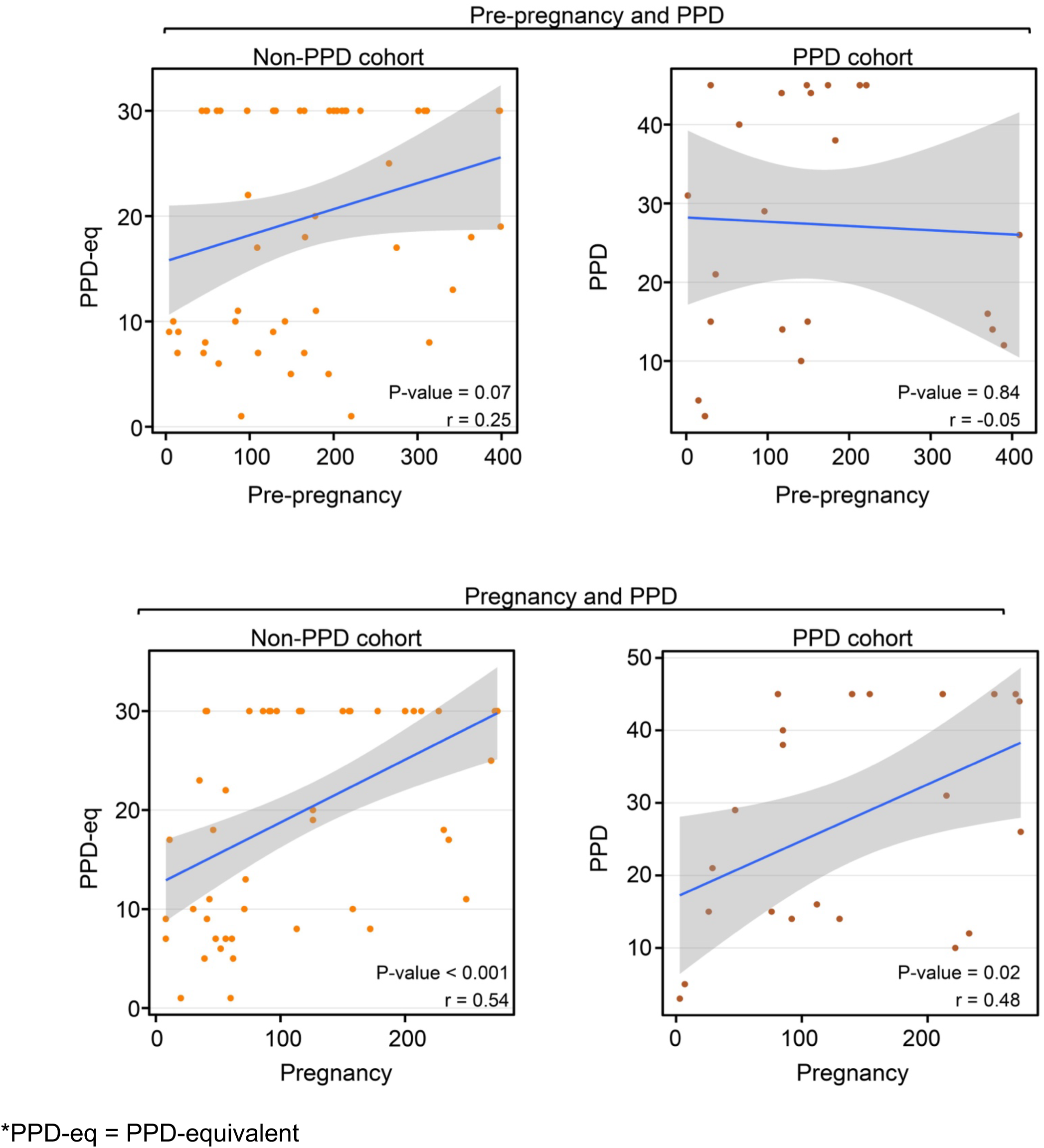
Consistent Fitbit wear time during pre-pregnancy was not correlated with PPD wear time consistency among women in the PPD cohort. The Pearson correlation coefficient between the maximum number of days in a row women PPD and non-PPD cohorts wore their Fitbit during pre-pregnancy and PPD (top) in addition to pregnancy and PPD (bottom). The blue line represents the line of best fit and gray shading shows the 95% confidence interval.

## Discussion

Our study elucidated numerous insights related to Fitbit wear time and PPD across time periods of pregnancy. First, we observed a strong trend that women with PPD wear their device a higher percentage of days than those without PPD during the postpartum and PPD time periods (Figure 2). However, among the days that women in the PPD and non-PPD cohorts wore their device, there was not a significantly different number of daily hours the device was worn (Figure 3). In terms of Fitbit wear time behavior to sleep, a similar pattern to percentage of daily wear time was observed with women in the PPD cohort wearing their Fitbit to sleep a higher percentage of days during the postpartum and PPD time periods compared to those in the non-PPD cohort (Figure 4). It was also seen that women in the PPD and non-PPD cohorts displayed the same correlation patterns between wear time during earlier and later pregnancy time periods (Figure 5). Finally, we found that women in both the PPD and non-PPD cohorts who wore their devices more consistently during pregnancy also maintained higher levels of device wear consistency during the PPD (or PPD-equivalent) periods. However, there was no correlation in the consistency of Fitbit wear time during the pre-pregnancy and PPD time periods among women in the PPD cohort (Figure 6).

Our study’s first key finding showed that women with PPD wore their Fitbit a higher percentage of days compared to women without PPD. A similar trend was detected during the postpartum time periods amongst women in the PPD and non-PPD cohorts (Figure 2). One reason we hypothesize that women with PPD wore their devices more frequently than those without PPD is due to anxiety and hypervigilance, which commonly occurs in women with PPD, and may drive increased personal tracking behavior^42^. Since PPD often goes undetected, women in the PPD cohort during the postpartum period (prior to EHR diagnosis) may have already been experiencing PPD symptoms, which could explain the similarities in patterns observed between the postpartum and PPD periods^4^. Unfortunately, AoURP does not have symptom-related data so we cannot know for sure when symptoms began. While it was originally suggested that PPD symptoms peak between 4-6 weeks in the postpartum period^43,44^, recent work suggests subgroups of women display unique symptom trajectories^45^. Further, although women in the PPD cohort tended to wear their Fitbit a higher percentage of days during the postpartum and PPD time periods, we did not detect any difference in the number of hours per day the device was worn compared to women without PPD (Figure 3). Our findings showed that both the PPD and non-PPD cohorts wore their devices approximately 15-17 hours of the day, which is consistent with other studies involving Fitbits^46,47^.

The next component of our study was to investigate the percentage of days women with PPD wear their device to sleep across each phase of pregnancy compared to those without PPD given the extensive relationship between sleep and PPD^35–41^. Our findings revealed women in the PPD cohort tended to wear their device more to sleep during the postpartum and PPD time periods compared to those without PPD (Figure 4). Considering the similar pattern observed in Fitbit wear time frequency (Figure 2), it was not surprising to find the same result in sleep data. Unfortunately, it is not possible to determine whether the device was intentionally worn for sleep tracking or simply due to continued use, but it could be interesting to investigate in future studies. The fact that women on average wear their device between 15-17 hours per day suggests that when women in these cohorts do wear their device, they also wear it to sleep^46,47^.

Lastly, our study sought to assess whether Fitbit wear time behavior during time periods prior to PPD may correlate with behavior during PPD, with the potential that wear time behavior during pre-pregnancy or pregnancy time periods may be able to help predict PPD onset. Our findings displayed that women who wear their device more during pre-pregnancy also wore their device more during PPD (Figure 5). A similar observation was detected when comparing pregnancy and PPD time periods; however, this relationship also persisted among women in the non-PPD cohort (Figure 5). When assessing the consistency of Fitbit wear time, we noticed a strong trend only in women without PPD that greater wear time consistency in pre-pregnancy correlated with greater consistency during the PPD-equivalent period (Figure 6). This may be attributed to women with PPD experiencing co-occurring mood and anxiety symptoms, leading them to wear their devices more frequently regardless of pre-pregnancy consistency^42^.

While this study is the first to evaluate wearable device wear time behavior amongst women with PPD across phases of pregnancy, it is not without limitations. First, the number of hours the device was worn was estimated based on recorded Fitbit steps data using previously established methods; however, it is not “ground truth” data and therefore may contain some level of inaccuracy^29^. Second, we do not have access to study participants in AoURP to perform any qualitative analysis to further understand causal relationships about individual-level Fitbit wear time patterns and disease symptoms/severity. Future studies should include user-experience-related questionnaires and qualitative methods tailored towards women with and without PPD during the postpartum period to assess the connection between PPD, hypervigilance, and device wear time^15–19^. Third, this study only investigated wearable device behavior for the Fitbit. While the Fitbit is the most commonly wearable device used for research purposes, it would be valuable to incorporate women with other devices, such as the Apple Watch, Google Fit, Garmin smartwatch, or Oura ring, which has shown high levels of adherence, and the type of device could be adjusted as a covariate^48–51^. Fourth, the PPD and non-PPD cohorts were relatively small and we posit we may have observed statistical significance with larger sample sizes. Finally, these cohorts consisted primarily of women who were white and non-Hispanic, where results may not generalize to other patient populations. Future work should involve more diverse populations to validate our findings and evaluate differences in wear time behavior between PPD and non-PPD cohorts across different racial/ethnic groups. Notably, one strength of this study is that AoURP does not send any type of notification or reminders for continued usage, thus our work provides a great foundation for the first study to assess real-world wearable device wear time behavior in women with PPD.

Understanding wearable device wear time behavior can provide insightful clinical information related to women with PPD. Considering that screening and diagnosis pose significant challenges, wearables, including features of wear time behavior, could potentially offer a viable solution. We envision a future using wearables combined with a machine learning algorithm that incorporates wearable device wear time and other digital biomarkers like sleep and physical activity, facilitate early detection of PPD by notifying the clinician with potential concern to prompt timely screening. Wear time behavior presents a passive and relatively straightforward feature to aid in evaluating PPD in non-clinical environments, and its application could potentially extend to other perinatal and general mental health disorders.

## Acknowledgements

First and foremost, the *All of Us* Research Program would not be possible without the partnership of its participants. The *All of Us* Research Program is supported by the National Institutes of Health, Office of the Director: Regional Medical Centers: 1 OT2 OD026549; 1 OT2 OD026554; 1 OT2 OD026557; 1 OT2 OD026556; 1 OT2 OD026550; 1 OT2 OD 026552; 1 OT2 OD026553; 1 OT2 OD026548; 1 OT2 OD026551; 1 OT2 OD026555; IAA #: AOD 16037; Federally Qualified Health Centers: HHSN 263201600085U; Data and Research Center: 5 U2C OD023196; Biobank: 1 U24 OD023121; The Participant Center: U24 OD023176; Participant Technology Systems Center: 1 U24 OD023163; Communications and Engagement: 3 OT2 OD023205; 3 OT2 OD023206; and Community Partners: 1 OT2 OD025277; 3 OT2 OD025315; 1 OT2 OD025337; 1 OT2 OD025276. We would also like to acknowledge that we greatly appreciate support from UNC Department of Genetics and this work would not have been possible without them.

## Competing interests

MAH is a founder of Alamya Health.

## Funding

Individual authors were supported by the following funding sources: NIMH R01131542 (RCP), NHGRI U24HG011449 (EH and MAH), NHGRI RM1HG010860 (EH and MAH).

## Author contributions

EH conceived the study. EH and MAH designed the study. EH analyzed the data. EH, SM, ZB, RCP, NE, and MAH interpreted the results. EH wrote the draft manuscript. MAH provided supervision and acquired funding. All authors reviewed and edited the final manuscript for publication.

## Data availability

Individuals who successfully complete the requisite training can access the data utilized in this analysis via the *All of Us* Researcher Workbench.

## Notes

### Author Declarations

All of Us Institutional Review Board gave ethical approval for this work.

## References

1. Insel, T. R. Digital Phenotyping: Technology for a New Science of Behavior. JAMA 318, 1215– 1216 (2017).10.1097/HRP.0000000000000013

2. Dunn, J., Runge, R. & Snyder, M. Wearables and the medical revolution. Pers. Med. 15, 429–448 (2018).

3. Gaynes, B. N. et al. Perinatal depression: prevalence, screening accuracy, and screening outcomes. Evid. Rep. Technol. Assess. (Summ.) 1–8 (2005) doi:10.1037/e439372005-001.

4. Cox, E. Q., Sowa, N. A., Meltzer-Brody, S. E. & Gaynes, B. N. The Perinatal Depression Treatment Cascade: Baby Steps Toward Improving Outcomes. J. Clin. Psychiatry 77, 20901 (2016).

5. Kingston, D., Tough, S. & Whitfield, H. Prenatal and Postpartum Maternal Psychological Distress and Infant Development: A Systematic Review. Child Psychiatry Hum. Dev. 43, 683–714 (2012).

6. Luca, D. L. et al. Financial Toll of Untreated Perinatal Mood and Anxiety Disorders Among 2017 Births in the United States. Am. J. Public Health 110, 888–896 (2020).

7. Reid, H. E., Pratt, D., Edge, D. & Wittkowski, A. Maternal Suicide Ideation and Behaviour During Pregnancy and the First Postpartum Year: A Systematic Review of Psychological and Psychosocial Risk Factors. Front. Psychiatry 13, 765118 (2022).

8. Postpartum Depression: Action Towards Causes and Treatment (PACT) Consortium. Heterogeneity of postpartum depression: a latent class analysis. Lancet Psychiatry 2, 59–67 (2015).

9. Guedeney, N., Fermanian, J., Guelfi, J. D. & Kumar, R. C. The Edinburgh Postnatal Depression Scale (EPDS) and the detection of major depressive disorders in early postpartum: some concerns about false negatives. J. Affect. Disord. 61, 107–112 (2000).

10. Deligiannidis, K. M. et al. Zuranolone for the Treatment of Postpartum Depression. Am. J. Psychiatry 180, 668–675 (2023).

11. Meltzer-Brody, S. et al. Brexanolone injection in post-partum depression: two multicentre, double-blind, randomised, placebo-controlled, phase 3 trials. The Lancet 392, 1058–1070 (2018).

12. Hurwitz, E. et al. Harnessing Consumer Wearable Digital Biomarkers for Individualized Recognition of Postpartum Depression Using the All of Us Research Program Data Set: Cross-Sectional Study. JMIR MHealth UHealth 12, e54622 (2024).

13. Ravindra, N. G. et al. Deep representation learning identifies associations between physical activity and sleep patterns during pregnancy and prematurity. Npj Digit. Med. 6, 1–16 (2023).

14. Sarhaddi, F., et al. Maternal Social Loneliness Detection Using Passive Sensing Through Continuous Monitoring in Everyday Settings: Longitudinal Study. JMIR Form. Res. 7, e47950 (2023).

15. Jeong, H., Kim, H., Kim, R., Lee, U. & Jeong, Y. Smartwatch Wearing Behavior Analysis: A Longitudinal Study. Proc. ACM Interact. Mob. Wearable Ubiquitous Technol. 1, 60:1–60:31 (2017).

16. Ogbanufe, O. & Gerhart, N. Watch It! Factors Driving Continued Feature Use of the Smartwatch. Int. J. Human–Computer Interact. 34, 999–1014 (2018).

17. Saheb, T., Cabanillas, F. J. L. & Higueras, E. The risks and benefits of Internet of Things (IoT) and their influence on smartwatch use. Span. J. Mark. - ESIC 26, 309–324 (2022).

18. Siepmann, C. & Kowalczuk, P. Understanding continued smartwatch usage: the role of emotional as well as health and fitness factors. Electron. Mark. 31, 795–809 (2021).

19. Visuri, A. et al. Understanding usage style transformation during long-term smartwatch use. Pers. Ubiquitous Comput. 25, 535–549 (2021).

20. Pathiravasan, C. H. et al. Factors associated with long-term use of digital devices in the electronic Framingham Heart Study. Npj Digit. Med. 5, 1–11 (2022).

21. All of Us Research Program Investigators. The “All of Us” Research Program. N. Engl. J. Med. 381, 668–676 (2019).

22. Holko, M. et al. Wearable fitness tracker use in federally qualified health center patients: strategies to improve the health of all of us using digital health devices. Npj Digit. Med. 5, 1–6 (2022).

23. Jooma, S. (NIH/OD) [E]. Data and Statistics Dissemination Policy. (2020).

24. Jones, S. E. et al. Who is pregnant? Defining real-world data-based pregnancy episodes in the National COVID Cohort Collaborative (N3C). JAMIA Open 6, ooad067 (2023).

25. Goodman, J. H. Postpartum depression beyond the early postpartum period. J. Obstet. Gynecol. Neonatal Nurs. JOGNN 33, 410–420 (2004).

26. Horowitz, J. A. & Goodman, J. A longitudinal study of maternal postpartum depression symptoms. Res. Theory Nurs. Pract. 18, 149–163 (2004).

27. Machado-Vieira, R. et al. The Timing of Antidepressant Effects: A Comparison of Diverse Pharmacological and Somatic Treatments. Pharm. Basel Switz. 3, 19–41 (2010).

28. Mughal, S., Azhar, Y. & Siddiqui, W. Postpartum Depression. in StatPearls (StatPearls Publishing, Treasure Island (FL), 2023).

29. Master, H. et al. Association of step counts over time with the risk of chronic disease in the All of Us Research Program. Nat. Med. 28, 2301–2308 (2022).

30. Lenth, R. emmeans: Estimated Marginal Means, aka Least-Squares Means. https://rvlenth.github.io/emmeans/ (2024).

31. Bates, D., Mächler, M., Bolker, B. & Walker, S. Fitting Linear Mixed-Effects Models Using lme4. J. Stat. Softw. 67, 1–48 (2015).

32. Kuznetsova, A., Brockhoff, P. B. & Christensen, R. H. B. lmerTest Package: Tests in Linear Mixed Effects Models. J. Stat. Softw. 82, 1–26 (2017).

33. Kassambara, A. ggpubr: ‘ggplot2’ Based Publication Ready Plots. (2023).

34. Benesty, J., Chen, J., Huang, Y. & Cohen, I. Pearson Correlation Coefficient. in Noise Reduction in Speech Processing (eds. Cohen, I., Huang, Y., Chen, J. & Benesty, J.) 1–4 (Springer, Berlin, Heidelberg, 2009). doi:10.1007/978-3-642-00296-0_5.

35. Iranpour, S., Kheirabadi, G. R., Esmaillzadeh, A., Heidari-Beni, M. & Maracy, M. R. Association between sleep quality and postpartum depression. J. Res. Med. Sci. 21, 110 (2016).

36. Lawson, A., Murphy, K. E., Sloan, E., Uleryk, E. & Dalfen, A. The relationship between sleep and postpartum mental disorders: A systematic review. J. Affect. Disord. 176, 65–77 (2015).

37. Lewis, B. A., Gjerdingen, D., Schuver, K., Avery, M. & Marcus, B. H. The effect of sleep pattern changes on postpartum depressive symptoms. BMC Womens Health 18, 12 (2018).

38. McEvoy, K. M. et al. Poor Postpartum Sleep Quality Predicts Subsequent Postpartum Depressive Symptoms in a High-Risk Sample. J. Clin. Sleep Med. JCSM Off. Publ. Am. Acad. Sleep Med. 15, 1303–1310 (2019).

39. Okun, M. L. Disturbed Sleep and Postpartum Depression. Curr. Psychiatry Rep. 18, 66 (2016).

40. Park, E. M., Meltzer-Brody, S. & Stickgold, R. Poor sleep maintenance and subjective sleep quality are associated with postpartum maternal depression symptom severity. Arch. Womens Ment. Health 16, 539–547 (2013).

41. Posmontier, B. Sleep Quality in Women With and Without Postpartum Depression. J. Obstet. Gynecol. Neonatal Nurs. 37, 722–737 (2008).

42. Thurgood, S., Avery, D. M. & Williamson, L. Postpartum Depression (PPD). Postpartum Depress. (2009).

43. Haga, S. M. et al. A longitudinal study of postpartum depressive symptoms: multilevel growth curve analyses of emotion regulation strategies, breastfeeding self-efficacy, and social support. Arch. Womens Ment. Health 15, 175–184 (2012).

44. Vliegen, N., Casalin, S. & Luyten, P. The Course of Postpartum Depression: A Review of Longitudinal Studies. Harv. Rev. Psychiatry 22, 1 (2014).

45. Drozd, F., Haga, S. M., Valla, L. & Slinning, K. Latent trajectory classes of postpartum depressive symptoms: A regional population-based longitudinal study. J. Affect. Disord. 241, 29–36 (2018).

46. Liao, Y. et al. Investigating the within-person relationships between activity levels and sleep duration using Fitbit data. Transl. Behav. Med. 11, 619–624 (2021).

47. Singh, B., Spence, R. R., Sandler, C. X., Tanner, J. & Hayes, S. C. Feasibility and effect of a physical activity counselling session with or without provision of an activity tracker on maintenance of physical activity in women with breast cancer — A randomised controlled trial. J. Sci. Med. Sport 23, 283–290 (2020).

48. Browne, J. D. et al. Lifestyle Modification Using a Wearable Biometric Ring and Guided Feedback Improve Sleep and Exercise Behaviors: A 12-Month Randomized, Placebo-Controlled Study. Front. Physiol. 12, (2021).

49. Care Evolution. Wearable devices | White Paper. CareEvolution https://careevolution.com/white-papers/choosing-a-commercially-available-wearable-device-for-your-research-study/ (2023).

50. Nolasco, H. R., Vargo, A., Bohley, N., Brinkhaus, C. & Kise, K. Examining Participant Adherence with Wearables in an In-the-Wild Setting. Sensors 23, 6479 (2023).

51. Shiba, S. K. et al. Assessing Adherence to Multi-Modal Oura Ring Wearables From COVID-19 Detection Among Healthcare Workers. Cureus (2023) doi:10.7759/cureus.45362.

